# Assessment of medication Dosage Adjustment in Hospitalized Patients with Chronic Kidney Disease

**DOI:** 10.1101/2020.05.04.20090787

**Authors:** Zair Hassan, Iftikhar Ali, Arslan Rahat Ullah, Raheel Ahmed, Shakeel Rehman, Azizullah Khan

## Abstract

**Background:** Inappropriate medication dosing can develop adverse drug reactions (ADRs) or ineffective therapy due to declined renal function in patients with renal insufficiency. This necessitates proper renal dose adjustment. Using a retrospective analysis of medical records, this study was proposed to evaluate medication dosage adjustment in hospitalized chronic kidney disease (CKD) patients.

**Methods:** This retrospective review of medical records was conducted at the Institute of Kidney Disease (IKD), Peshawar. It included all CKD patients hospitalized between May 01, 2019 and April 25, 2020. Glomerular filtration rate was calculated using Modification of Diet in Renal Disease (MDRD) formula and dose appropriateness was established by evaluating practice with relevant reference books.

**Results:** Of the total 1537 CKD patients, 231(15.03%) had evidence of dosing error, which were considered for final analysis. Overall, 1549 drugs were prescribed, 480(30.99%) drugs required dose adjustment of which 196(40.42%) were adjusted properly and the remaining 286(59.58%) were unadjusted. The most common unadjusted drugs were meropenem, cefepime, ciprofloxacin and rosuvastatin, whereas captopril, aspirin, bisoprolol, pregabalin and levofloxacin had the highest percentage of adjusted drugs. On multivariate logistic regression, the number of drugs requiring dosing adjustments and obstructive nephropathy were found to be statistically significant factors that increased the likelihood of the medication dosing errors: A unit increase in the number of drugs requiring dose adjustment increases 5.241 times the likelihood of dosing error. Similarly the presence of obstructive nephropathy (OR 0.383, 95% Cl [0.153-0.960] P= 0.041) was found to be significantly associated with dosing error after adjustment for potential confounding factors.

**Conclusion:** The dosing of more than half of the prescribed drugs that required adjustment in CKD patients were not adjusted which showed that medication dosing errors were high. This highlights the importance of medication prescription according to guidelines in these patients to improve the outcomes of pharmacotherapy and patients’ quality of life.

## INTRODUCTION

Chronic kidney disease (CKD) is defined as “a decrease in glomerular filtration rate (GFR) <60mL/minute /1.73m^2^ for ≥3 months”, irrespective of the etiology and is classified into five stages based on GFR. CKD affects 8%–16% of the people globally, and is one of the major public health challenges. ^1, 2^ An approximately 5 to 10 million deaths are attributed to kidney diseases every year.^3^ The prevalence of CKD in Pakistan is high, affecting 12.5% (11.4%–13.8%) of the population.^4^

Kidneys are key structures responsible for regulating homeostasis, acid-base equilibrium, as well as electrolytes. ^5^ Most of the drug’s metabolism and removal depend on normal kidney function.^6^ Maintaining healthy kidney function is indeed necessary for several drugs and potential active metabolites to be metabolized and eliminated. Unlike in patients with healthy kidney function, the recommended dose of medications in patients with renal impairment can cause adverse drug reactions owing to impaired kidney function of patients.^5-8^ Furthermore, to minimize adverse drug reactions and therapeutic failure, dosage modifications according to renal function in patients with renal failure need to be individualized.

CKD patients are taking number of medications not only for preventing the progression of the disease process, but also for comorbidities. With a decrease in renal function, the pharmacokinetic parameters of several drugs are so ominously changed that the normal doses transform either to augmented or diminished. ^9-11^ In admitted patients, lethal or ineffective doses may increase the hospital stay, treatment cost, and accordingly, adding extra burden on both patient and the healthcare systems.

In CKD patients, the kidney’s ability to eliminate a drug is compromised resulting in accumulation of drugs, increasing the adverse effects and possibly leading to toxicities/ adverse drug events (ADEs).^7^ Renal injury is a known risk factor for ADEs, but remains ignored very often by Healthcare professionals.^12,13^ A study by Hug BL et al revealed that 10% of patients with CKD experienced an ADE, out of these, 91% were considered preventable and 51% were serious. ^14^ A number of published articles have described considerable dose adjustment related difficulties and medication errors in CKD patients.^14-16^ Poor understanding of the importance and optimization of medication dosages is often a cause of prescribing errors in patients with compromised renal function. ^15, 17^

The most common dosing error while managing CKD patients is those observed during antimicrobial use, requiring a lookout and adjustment in these patients depending on eGFR of patients.^18^ Studies from China, showed antibiotics related dosage error in CKD patients were 38.85%–60.3%.^19, 20^

Literature support that medications that required dose adjustment in CKD patients are not adjusted accurately in admitted patients and the practice is common in both developed and developing countries; during hospitalization about 25%- 77% drugs are adjusted inappropriately.^13,21^ In developed countries like Netherlands a study on CKD patients with advance stages (stage III and IV) reveals a high prevalence of unadjusted dose prescription.^22^

Similarly, a study in Australia also reported a high level of inappropriate prescription in elderly CKD patients with diabetes, with a polypharmacy.^23^ To address such an important problem, multiple research studies have been published globally to evaluate the dosing errors pattern; although the subject is not thoroughly investigated particularly in the developing countries.

A literature search showed very limited published material on this subject in Pakistan. As of now, no published studies were to our knowledge carried out in this part of the country (Peshawar). Considering this, the study was proposed to evaluate medication dosage adjustment in admitted CKD patients in Peshawar, Pakistan.

## METHODS

### Study Setting

This retrospective study was conducted from May 01, 2019 and April 25, 2020 at Nephrology department of Institute of Kidney Disease (IKD), Peshawar, Pakistan. The Institute is a tertiary care health facility in Peshawar, the capital of Khyber Pakhtunkhwa.

### Study Design and Sampling Procedure

Data was extracted from medical record/patients files of CKD patients admitted to nephrology ward on structured format. All diagnosed cases of CKD receiving at least one drug requiring adjustment with length of hospital stay greater than 24 hours were included. The GFR was calculated using “Modification of Diet in Renal Disease (MDRD)”.^24^ Cockcroft-Gault formula^25^ was not applied to calculate GFR due to missing of patient’s body weight. The GFR stage was determined for individual patient depending on his current condition agreeing to “Kidney disease: improving global outcomes” (KDIGO 2012) guidelines. The results of identified patients belonged to the GFR stages G3a (45-59 ml/min), G3b (30-44 ml/min), G4 (15-29 ml/min), and G5 (<15 ml/min).

### Assessment of Medication Dosing Errors

Unfortunately, due to non-availability of any national drug formulary and drug dosing guidelines for CKD patients in Pakistan, we had to rely on some of the reputed references and dose adjustment guidelines for the individual drug doses assessment for appropriateness by comparing the observed practices.

Dosage appropriateness was based on comparing the practice with the established recommendation: “Drug Dosing Adjustments in Patients with Chronic Kidney Disease” published by the American Academy of Family Physicians, and “Drug Information Handbook, 25^th^ edition” published by Lexicomp®.^26^ The guidelines were selected in consultation with consultant Nephrologist.

The dose appropriateness was evaluated by a physician and hospital pharmacist. The prescribed doses which were in accordance with the recommended guidelines were regarded as adjusted. Nonetheless, inappropriately dosed or dose given recommended for patients with normal renal function was categorized as not adjusted.

### Data Analysis

SPSS Version 20 was used for data analysis. Descriptive statistics were used to present the results such as frequency and percentage for categorical data while mean (SD) or Median (IQR) where appropriate for numerical data. Logistic regression models were used to calculate the associated factors “taking all medications per patient” were not adjusted [(Yes/No)] as the main outcome measure that describes the medication error. Results were described as Odd ratios (OR) along with 95%Cl. Those variables (p <0.2) in the univariate analysis were included in the multivariate statistics. P < 0.05 was measured statistically significant.

### Ethical Considerations

The research protocol was reviewed and approved by Institutional Ethical Review Committee of Hayatabad Medical Complex-Peshawar, Pakistan (Ref number: 1510-2019).

### Results

During the study period, a total of 1537 CKD patients’ medical record/charts were reviewed. However, a total of 231 patients (15.03% of screened patients) were included in the final analysis. The demographics and clinical characteristics of patients are listed in ***table 01***. Of the total 231 patients, 184(79.7%) were males and 47(20.34%) were females. The mean (SD) age of the patients was 46.14±15.90 years (Range: 13-85 years). The mean (SD) length of the hospital stay was 3.97±1.96 days (Range: 02-16 days). Majority of the patients 209 (90.5%) were in G5 stage of CKD, followed by 14(6.1%) in G4. The mean (SD) of the drugs prescribed were 6.7±1.33. About 85.3% patients were prescribed >5 drugs. Similarly majority of the patients 220(95.23%) medications list were comprised of antibiotics. Comorbidities were present in most of the patients 180(77.92%). Among the comorbid conditions hypertension 148(64%), diabetes mellitus 57(24.67%) and obstructive nephropathy 36(15.58%) were on the top of the list.

**Table. 1.** Demographic and clinical characteristics of the patients (N=231)

|  |  |
| --- | --- |
| **Variable** | **N (%)** |
| **Age (years)**[mean (SD)] | 46.14±15.90 |
| **Gender** |  |
| Male | 184(79.65) |
| Female | 47(20.35) |
| **Length of hospital stay (days)** [mean (SD)] | 3.97±1.96 |
| **GFR category** |  |
| G3b | 08(3.46) |
| G4 | 14(6.06) |
| G5 | 209(90.48) |
| **eGFR** ml/min./1.73m<sup>2</sup> [mean (SD)] | 8.07±7.8 |
| **BUN** mg/dl[mean ±SD] | 205.29±93.02 |
| **Serum Creatinine** mg/dl[mean ±SD] | 10.33±5.43 |
| **K** mmol/l[mean ±SD] | 4.89±1.01 |
| **Number of drug prescribed**[mean ±SD] | 6.7±1.33 |
| ≤5 | 34(14.72) |
| >5 | 197(85.28) |
| **Antibiotics prescribed** |  |
| Yes | 220(95.24) |
| No | 11(4.76) |
| **Comorbidities present** |  |
| Yes | 180(77.92) |
| No | 51(22.07) |
| **Diabetes mellitus** |  |
| Yes | 57(24.68) |
| No | 174(75.32) |
| **Hypertension** |  |
| Yes | 148(64.07) |
| No | 43(18.61) |
| **Ischemic heart disease** |  |
| Yes | 21(9.09) |
| No | 210(90.91) |
| **Urinary tract infection** |  |
| Yes | 19(8.23) |
| No | 212(91.77) |
| **Hepatitis B** |  |
| Yes | 09(3.90) |
| No | 222(96.10) |
| **Hepatitis C** |  |
| Yes | 21(9.09) |
| No | 210(90.91) |
| **Obstructive nephropathy** |  |
| Yes | 36(15.58) |
| No | 195(84.42) |

A total of 1549 numbers of drugs were prescribed in the study patients. Of the total prescribed drugs 480 (30.99%) required dosage adjustment. Among them, 286 (59.58%) were unadjusted, and the remaining 194(40.42%) were properly adjusted as depicted in ***table 02***.

**Table. 2.** Number and mean or median of properly adjusted and unadjusted drugs prescribed

|  |  |  |  |  |
| --- | --- | --- | --- | --- |
| ***Variable*** | ***N*** | ***%*** | ***Mean(SD)*** | ***Median (IQR)*** |
| Total number of drugs prescribed | 1549 | 100 | 6.7±1.33 |  |
| Total number of drug prescribed requiring adjustment | 480/1549 | 30.99 | 2.08±0.86 |  |
| Total number of drugs properly adjusted | 194/480 | 40.42 |  | 1(0-3) |
| Number of drugs unadjusted | 286/480 | 59.58 |  | 1(0-3) |

The descriptive statistics as depicted in ***table 3*** showed that the most frequently unadjusted drugs were Meropenem (100%), Cefepime (100%), Ciprofloxacin (100%), Rosuvastatin (100%), Cefoperazone/sulbactam (91.33%), Ranitidine (65.71%) and Piperacillin/Tazobactam (85.71%), while in comparison the most accurate properly adjusted drugs were aspirin (100%), captopril (100%), bisoprolol (100%), Pregabalin (100%), levofloxacin (100%), vancomycin (87.5%), domiperidone (80.7%), Cefotaxime (78.12%), furosemide (69%), sodium bicarbonate (53.65%) and spironolactone (50%).

**Table. 3.** List of drugs needing adjustment, properly adjusted, or unadjusted

|  |  |  |  |
| --- | --- | --- | --- |
| **Drug name** | **Drugs needing adjustment (N)** | **Drugs adjusted N (%)** | **Drugs unadjusted N (%)** |
| Meropenem | 17 | - | 17(100) |
| Sodium Bicarbonate | 41 | 22(53.66) | 19(46.34) |
| Domiperidone | 52 | 42(80.77) | 10(19.23) |
| Cefoperazone/sulbactam | 127 | 11(8.66) | 116(91.34) |
| Ranitidine | 70 | 24(34.29) | 46(65.71) |
| Furosemide | 55 | 38(69.09) | 17(30.91) |
| Cefotaxime | 32 | 25(78.12) | 7(21.87) |
| Metoclopramide | 6 | 1(16.66) | 5(83.33) |
| Piperacillin/tazobactam | 7 | 1(14.28) | 6(85.71) |
| Tranxemic acid | 3 | 1(33.33) | 2(66.66) |
| Vancomycin | 8 | 7(87.5) | 1(12.5) |
| Captopril | 3 | 3(100) | - |
| Aspirin | 6 | 6(100) | - |
| Ciprofloxacin | 3 | - | 3(100) |
| Cefepime | 27 | - | 27(100) |
| Spironolactone | 6 | 3(50) | 3(50) |
| Rosuvastatin | 3 | - | 3(100) |
| Ramipril | 2 | 1(50) | 1(50) |
| Bisoprolol | 2 | 2(100) | - |
| Fluconazole | 5 | 2(40) | 3(60) |
| Pregabalin | 2 | 2(100) | - |
| Levofloxacin | 2 | 2(100) | - |
| Linezolid | 1 | 1(100) | - |
|  | **480** | **194(40.42)** | **286(59.58)** |

Of the total variables, GFR category G5 (odds ratio OR 0.340, 95%Cl [0.121-0.955] P= 0.041), total number of drug prescribed (OR 1.826, 95%Cl [1.444-2.310] P≤0.01) and drug requiring dose adjustment (OR 4.818, 95%Cl [3.054-7.600] P<0.001) were noted to be significantly associated with dosing error on univariate analysis as shown in ***table 4***. Multivariate analysis was run for the variables that were found significant (P≤0.2) on univariate analysis. A unit increase in number of drugs requiring dose adjustment increases 5.241 times the likelihood of medication dosing error. Similarly, the presence of obstructive nephropathy (OR 0.383, 95%Cl [0.153-0.960] P= 0.041) was found to be significantly associated with medication error after adjustment for potential confounding factors.

**Table. 4:** Regression analysis of demographic and clinical variables with medication dosing errors

| **Variable** | **All medication per patient were un adjusted** |  | **Odds Ratio (cOR) 95%CI** | **p-value** |
| --- | --- | --- | --- | --- |
|  | **No** | **Yes** |  |  |
| **Age(years) [mean]** | 46.43 | 45.53 | 1.004[0.988-1.021] | 0.601 |
| **Gender** |  |  |  |  |
| Male | 100(77.5) | 84(82.4) | 0.739[0.384-1.424] | 0.366 |
| Female | 29(22.5) | 18(17.6) | Reference |  |
| **Length of hospital stay(days)[mean]** | 3.94 | 3.99 | 0.989[0.867-1.128] | 0.865 |
| **Serum Creatinine(mg/dl)** | 10.01 | 10.75 | 0.975[0.929-1.023] | 0.303 |
| **BUN(mg/dl)** | 204.68 | 206.06 | 1.000[0.997-1.003] | 0.911 |
| **K<sup>+</sup> (mmol/l)** | 4.91 | 4.87 | 1.038[0.802-1.344] | 0.777 |
| **GFR category** |  |  |  |  |
| G3b and G4 | 17(13.2) | 5(4.9) | Reference |  |
| G5 | 112(86.8) | 97(95.1) | 0.340[0.121-0.955] | 0.041 |
| **Total of drugs prescribed[mean]** | 7.12 | 6.18 | 1.826[1.444-2.310] | <0.001 |
| **Antibiotics prescribed** |  |  |  |  |
| No | 4(3.1) | 7(6.9) | Reference | 0.193 |
| Yes | 125(96.9) | 95(93.1) | 0.434[0.124-1.527] |  |
| **Drugs requiring dose adjustment[mean]** | 2.46 | 1.60 | 4.818[3.054-7.600] | <0.001 |
| **Comorbidity present** |  |  |  |  |
| Yes | 98(74.8) | 84(82.4) | Reference |  |
| No | 33(25.2) | 18(17.6) | 0.623[0.327-1.188] | 0.151 |
| **Diabetes mellitus** |  |  |  |  |
| Yes | 38(29.5) | 19(18.6) | 1.824[0.976-3.411] | 0.060 |
| No | 91(70.5) | 83(81.4) | Reference |  |
| **Hypertension** |  |  |  |  |
| Yes | 79(61.2) | 69(67.6) | 0.756[0.438-1.304] | 0.314 |
| No | 50(38.8) | 33(32.4) | Reference |  |
| **Ischemic heart disease** |  |  |  |  |
| Yes | 15(11.6) | 6(5.9) | 2.105[0.786-5.637] | 0.138 |
| No | 114(88.4) | 96(94.1) | Reference |  |
| **Urinary tract infection** |  |  |  |  |
| Yes | 10(7.8) | 9(8.8) | 0.868[0.339-2.224] | 0.769 |
| No | 119(92.2) | 93(91.2) | Reference |  |
| **Hepatitis C** |  |  |  |  |
| Yes | 11(8.5) | 10(9.8) | 0.858[0.349-2.107] | 0.738 |
| No | 118(91.5) | 92(90.2) | Reference |  |
| **Hepatitis B** |  |  |  |  |
| Yes | 4(3.1) | 5(4.9) | 0.621[0.162-2.374] | 0.486 |
| No | 125(96.9) | 97(95.1) | Reference |  |
| **Obstructive nephropathy** |  |  |  |  |
| Yes | 15(11.6) | 21(20.6) | 0.508[0.247-1.044] | 0.065 |
| No | 114(88.4) | 81(79.4) | Reference |  |

## DISCUSSION

CKD patients are the high-risk population for drug related problems, and among them, medication dosing errors are the most prevalent in these patients. ^7,27,28^ A large number of published studies ^7,15,19,27,34^ have determined the medication dosing error in these patients, but unfortunately, few to none published studies were, to our knowledge, conducted in this part of the region.

The economic burden of CKD and its associated complications is incurring the overall health care expenditures. Moreover, the early presentation of this chronic condition extends the burden to an even young population and in effect leading to further financial burdens.^3^ The Pakistani health care system is under resourced and overburdened, this along with organizational mismanagement [including lack of pharmacist in the multidisciplinary team and direct patient care] making it more worst. Keeping in view the existing health care system, there is likely the possibility of dosing error in these patients. The present study was proposed to assess medication dosing error in renal impaired hospitalized patients.

This study revealed that a total of 1549 drugs were prescribed to the patients with a mean [SD] 6.7±1.33. Of the total, 480(30.99%) medication orders needed dose adjustment. Whereof, about 40.42% were adjusted and the rest of (59.58%) were unadjusted almost similar to the finding of a study conducted in Bahawalpur, Pakistan by Ahsan Saleem and Imran Masood in CKD patients.^28^ Our findings are slightly higher than those reported by Saad et al in which 37% of the prescription order were adjusted adequately at two university hospitals in, Lebanon.^27^

In this study, the prevalence of medication dosing error was considerably lower as compared with previous reports from India (81%), Palestine (73.6%), South Africa (68%) and Lebanon (63%).^28,33,35^ The low percentage of medication error in these CKD patients might be due to the reason that they received treatment from trained nephrologist. In contrast to our study the result of medication dosing error was higher than even less developed countries such as Indonesia and Nepal where the unadjusted drugs percentage were 20.0% and 13.5% respectively.^29,32^ The proportion of dosing error in our study was also greater than the studies conducted in Saudi Arabia, Australia and France which is 53.1%, 44.8% and 34.0%, respectively.^17,30,31^ This suggests that either the physician’s knowledge is inadequate or the clinical pharmacy services in our setting are lacking, compared to developed states. In the less developed nations such as Indonesia and Nepal, the lower prevalence of medication dosing errors could be attributed to active involvement of clinical pharmacist in direct patient care. Whereas, Altunbas et al. reported considerably lower prevalence of medication dosing errors (12.6%), however the authors explained that majority of the study participants had renal impairment thereby necessitating nephrology consultation and optimized drug regimen, which could limit the generalizability of study findings.^36^

In developed nations, the active involvements of clinical pharmacists coupled with computerized dosage optimization systems are primarily responsible for the appropriate medication therapy. Most automated dosage adjustment systems automatically alert the healthcare professionals including physicians and clinical pharmacist regarding renal function status and the need for dose optimization.^17,33^ Therefore, lack of pharmacist in clinical setting, lack of national dosing formulary and computerized dose adjustment programs in Pakistan lead to higher medication dosing error.

The unadjusted drug proportion was higher in patients with G5, where 97/209 drug entries were unadjusted despite the fact that G5 is more advanced disease stage. A study conducted in Ethiopia reported that in patients with stage 5, 80% of the drugs were inappropriately adjusted.^37^ A study results by Saad et al described that drugs prescribed in patients with stage 5 nearly 37 % drug required dose adjustment.^27^

Another important aspect in this study is the unusual age distribution where the patient mean age is a 46.14±15.90 year which is much different from the developed and even under developing countries.^17,29,30^ This difference might be due to low expectancy of life in a normal healthy Pakistani which is almost 65 years, dying at an early age, before reaching the average life expectancy The high prevalence of CKD in young adult in Pakistan is due to high burden of diabetes mellitus, hypertension and renal stone, evident in our study result as well.^38,39^

While assessing the pattern of medication dosing error, the majority of prescribed drugs were ordered without consulting dose adjustment guideline. These include cephalosporin antibiotics (Cefoperazone/sulbactam, Cefotaxime, and Cefepime), Meropenem, Sodium bicarbonate, Ranitidine, Metoclopramide, Furosemide, Spironolactone, and Rosuvastatin. These findings are in agreement with earlier studies with the exception of Vancomycin and the cardiac medication that were prescribed more appropriately in our study sample. ^33, 37^ These observation shows that Pakistani Physicians in public healthcare facilities are underestimating the adverse events associated to several drugs. For instance, the antimicrobial like Aminoglycosides, Vancomycin and cephalosporins have the potential to induce nephrotoxicity.^37^

Considering, “unadjusted of all medication per patient” [yes or No] as dependent variable the results of logistic regression revealed that age, sex, length of hospital stay, antibiotics prescribed, Serum Creatinine, blood urea nitrogen, Potassium, diabetes mellitus, Hypertension, Ischemic heart disease, Urinary tract infection, Hepatitis B and C and obstructive nephropathy etc. did not show statistical significance. Similar findings have been reported by previous studies.^27,28,37^ However, GFR categories G5(cOR=0.340) had more unadjusted drugs compared to G3b and G4, whereas a unit increase in a drug prescription result an increase in unadjusted dose by a factor of 1.826. Similarly, total number of drugs per patient that required dose adjustment was significantly associated with medication error on univariate analysis. The findings of the present study are consistent with previous report by Ahsan Saleem and Imran^28^, in which, end stage CKD, and the number of prescribed drugs were significant determinants of medication dosing errors. In contrast, Getachew H et al. reported that severity of renal impairment, prescribed medications requiring dose optimization, and number of prescribed medications per patient differs with the percentage of properly adjusted drugs per patient.^37^

Medication dosing errors was not related to diabetes in this study, which is not in line with a study by Khanal A et al.^23^ Furthermore the presence of antimicrobial in patient prescription was not a significant predictor, similarly reported by Ahsan Saleem.^28^

Multivariate analysis showed that a unit increase in number of drugs requiring dose adjustment increases the chances of medication error by 5.241 times. Similarly the presence of obstructive nephropathy aOR 0.383 was found to be significantly associated with medication error after adjustment for potential confounding factors. Similar results have been described by a similar study^28^ where the numbers of prescribed medicines and presence of comorbidities were significant determinants of medication dosing errors.

In CKD patients the medications should be selected and prescribed with extreme precautions and appropriateness to avoid possible drug related problems and adverse outcomes. The predictors identified should be paid attention to.

### Strength and Limitations

This study envisages several strengths. The first strength among them is that it is the first study performed in tertiary care hospital of Peshawar Khyber Pakhtunkhwa and second in Pakistan. Secondly, in comparison to the previous study it is more detailed and to look into pattern and determinants of medication dosing errors identified in admitted CKD patients. Despite the strength the present study has several limitations. Firstly, the small sample size as compared to the whole CKD population. Secondly, being a retrospective study we could not intervene which restricted us from possibly suggesting intervention and observing actual adverse drug reaction. Thirdly, the MDRD formula was applied due to insufficient data on patient medical charts such as weight which is not suitable for higher muscle mass patients and those with malignant condition such as cancer. Fourthly, the nephrologist may have consulted dose adjustment guidelines other than we used. It is plausible to conclude that apart from renal function status, physicians may have based dosage optimization on blood pressure, serum electrolytes and heart rate. Finally, due to limited resources the present study was a single center data and finding cannot be generalized.

### Clinical Implications and Future Recommendations

An appraisal of the literature in correlation with the findings of the present study revealed that errors in medication dosing are a common clinical issue, particularly in patients with CKD. Dosing errors necessitate adequate attention by healthcare professionals (HCP) and CKD patients need to be specifically evaluated for dosage optimization before prescribing medication. Moreover, there exists considerable confusion regarding the need and extent of dosage adjustment in patients with varying degree of renal impairment, hence harmonized and universally acceptable guidelines should be formulated and adopted regarding the dosage optimization of renally excreted drugs so as to safeguard the patient’s health. In developing countries including Pakistan, the active involvement of trained clinical pharmacists in direct patient care should be ensured in order to promote appropriate and optimized medication therapy. Furthermore, continuous medical educational programs, seminars, and workshops need to be organized on a regular basis regarding the optimization of medication therapy, particularly in patients with renal impairment. Therefore, intensified collaboration between HCPs (general practitioners (GPs), nephrologists and clinical pharmacists) is recommended with relevant exchange of patient information in a bid to reducing the rate of inappropriate prescription and medication dosing errors.

## CONCLUSION

The medication related dosing errors was quite high, more than half of the drugs need dose adjustment. The result also showed that appropriate dose adjustment for impaired kidney function was not accomplished in a large number of patients in the province’s largest teaching hospital with specialist nephrologists and qualified residents who are expected to have a greater knowledge of dose adjustment in CKD patients compared to doctors in general hospitals. The determinants of medication associated dosing error in our study were the number of drugs requiring dose adjustment and obstructive nephropathy. This result suggests the need to provide physicians with dose adjustment information and recommendations in CKD patients in order to avoid dosing errors in such patients.

## Data Availability

The datasets generated and/or analyzed during the current study are not publicly available but are available from the Corresponding author on reasonable request.

## Acknowledgments

We would like to express our great appreciation to Beenish Mehmood, (Physical Therapist, Paraplegic Center, Peshawar) and Mohsin Zafar (Cardiology Resident, Lady reading hospital Peshawar) for their contribution, feedback and input.

## Author Contributions

Conception and design: ZH, IA and AR, U. Data collection: ZH, RA. Data analysis and interpretation, Results: IA, ZH. Manuscript drafting and writing: IA, ZH, SR. Language editing, appropriateness, critical revision: AR U, AZ, And IA. All authors read and approved the final version of the paper.

## Notes

### Competing Interest Statement

The authors have declared no competing interest.

### Funding Statement

The authors received no financial support for the research, authorship, and /or publication of this article.

